# Automated Prioritization of Sick Newborns for Whole Genome Sequencing Using Clinical Natural Language Processing and Machine Learning

**DOI:** 10.1101/2022.05.06.22274688

**Authors:** Bennet Peterson, Javier Hernandez, Charlotte Hobbs, Sabrina Malone Jenkins, Barry Moore, Edwin Juarez, Samuel Zoucha, Erica Sanford Kobayashi, Matthew N. Bainbridge, Albert Oriol, Luca Brunelli, Stephen Kingsmore, Mark Yandell

## Abstract

**Background:** Rapidly and efficiently identifying critically ill infants for WGS is a costly and challenging task currently performed by scarce, highly trained experts, and is a major bottleneck for application of WGS in the NICU. Automated means to prioritize patients for WGS are thus badly needed.

**Methods:** Institutional databases of Electronic Health Records (EHRs) are logical starting points for identifying patients with undiagnosed Mendelian diseases. We have developed automated means to prioritize patients for Rapid and Whole Genome Sequencing (rWGS and WGS) directly from clinical notes. Our approach combines a Clinical Natural Language Processing (CNLP) workflow with a machine learning-based prioritization tool we call *the Mendelian Phenotype Search Engine* (MPSE).

**Results:** MPSE accurately and robustly identified NICU patients selected for WGS by clinical experts from Rady Children’s Hospital in San Diego (AUC 0.86) and the University of Utah (AUC 0.85). In addition to effectively identifying patients for WGS, MPSE scores also strongly prioritize diagnostic cases over non-diagnostic cases, with projected diagnostic yields exceeding 50% throughout the first and second quartiles of score-ranked patients.

**Conclusions:** Our results indicate that an entirely automated pipeline for selecting acutely ill infants in neonatal intensive care units (NICU) for WGS can meet or exceed diagnostic yields obtained through current selection procedures, which require time-consuming manual review of clinical notes and histories by specialized personnel.

## Introduction

It is estimated that 7 million infants are born worldwide with genetic disorders each year ^1^. Admission to the Neonatal Intensive Care Unit (NICU) often provides the first opportunity for their diagnosis and treatment. Disease can progress rapidly in acutely ill infants, necessitating timely diagnosis in the hope of implementing personalized interventions that can decrease morbidity and mortality. Thus, rapid Whole Genome Sequencing (rWGS) is increasingly being used as a first line diagnostic test ^2,3^.

Current estimates suggest that around 18% of neonates admitted to the NICU harbor a Mendelian disease, and rWGS diagnostic rates in this population are over 35% ^4,5^. Rapidly and efficiently identifying infants for WGS is costly and challenging, as large NICUs often see more than 1000 admissions per year, and neonatal clinical histories evolve rapidly from the time of admission. Previous studies of rWGS in the NICU used inclusion criteria that limited enrollment to the first 96 hours ^3,5^ or 7 days ^6^ of admission or development of an abnormal response to standard therapy for an underlying condition, but these restrictions may miss the earliest opportunity to sequence a neonate. Minute-to-minute changes in laboratory results, diagnostic imaging, and clinical trajectory suggest that constant automated vigilance, as opposed to one or two isolated points in time, may be optimal to identify infants most likely to benefit from WGS. Done manually, this would be prohibitively time-consuming and costly. Automated means to prioritize patients for WGS are thus badly needed. Indeed, this is the principal motivation for the work described here.

Phenotype descriptions are crucial components of the WGS diagnostic process, and many tools exist for combining phenotypic terms with WGS data to prioritize disease-causing variants ^7–10^. Current best practice is to describe patient phenotypes using Human Phenotype Ontology (HPO) terms ^11^. These descriptions usually take the form of machine-readable phenotype term lists, an important prerequisite for automated analyses.

Care providers emphasize the importance of clinical notes for informing disease diagnosis, and HPO-based phenotype descriptions are generally compiled through manual review of these free text documents. Unfortunately, this is a time-consuming process that requires highly trained experts, and is a major bottleneck for application of WGS in the NICU ^12,13^.

Natural Language Processing (NLP) is a class of computational methods for generating structured data from unstructured free text. Recent work has begun to explore the utility of using Clinical Natural Language Processing technologies (CNLP) to automatically generate descriptions directly from clinical notes, with several groups demonstrating that rWGS diagnosis rates using CNLP derived descriptions can equal or exceed those obtained using manually compiled ones ^12,14^. This is a significant step towards scalability and automation. The ability to automatically survey all NICU admissions daily, for example, would mean that rWGS candidates could be ranked as part of an ever-evolving triage process based upon the latest contents of their EHRs.

Although the use of HPO descriptions for WGS-based Mendelian diagnosis is now established practice ^7–10,14^, the benefit of prioritization of patients for sequencing based on HPO terms is not known. To explore the feasibility of such an approach, we have combined a CNLP workflow with a machine learning-based prioritization tool we call the Mendelian Phenotype Search Engine (MPSE). MPSE employs HPO-based phenotype descriptions derived from patient EHRs to compute a score. This score can be used to determine the likelihood that a Mendelian condition is contributing to a patient’s clinical presentation, and thus, can be used for the prioritization of patients for WGS. To demonstrate feasibility, we used a highly curated clinical dataset consisting of 1049 patients admitted to a Level IV NICU (the highest level of acuity for a NICU) and their clinic notes; 293 of these children had rWGS, with 85 receiving a diagnosis. Our cross validated results indicate that an entirely automated CNLP/MPSE-based selection process for rWGS can obtain diagnostic rates equaling or exceeding those obtained though manual review and selection as per current best practice. A second independent replication study at the University of Utah provides additional support for these conclusions, demonstrating that MPSE operates effectively at both institutions.

## Methods

### Datasets

Our clinical dataset consisted of 293 probands who underwent rWGS at Rady Children’s Hospital in San Diego (RCHSD), 85 of which received a molecular diagnosis of Mendelian disorder. The diagnosed individuals represent a real-world population comprised of different Mendelian conditions resulting from diverse modes of disease inheritance and disease-causing genotypes ^3,5,14^. To this cohort, we added every NICU admission at RCHSD in the year 2018. The 756 additional patients and their clinic notes provide a diversity of phenotypes not necessarily associated with Mendelian diseases. In total, the RCHSD dataset consisted of 1049 individuals. A second independent dataset of 35 probands that were sequenced as part of the University of Utah NeoSeq program ^15^ and 2930 randomly selected University of Utah Level-III NICU patients from 2010 to 2022, was retrospectively analyzed to evaluate the utility of the RCHSD training data for prioritizing probands for rWGS at a second institution.

### Phenotype descriptions

Highly-curated, manually created HPO-based phenotype descriptions were provided for each of the 293 RCHSD and 35 University of Utah WGS cases, as described in NSIGHT1 ^3^. Corresponding CNLP-derived phenotype descriptions were generated for all 1049 RCHSD probands and 2965 University of Utah probands by NLP analysis of clinical notes using CLiX ENRICH (Clinithink, Alpharetta, GA) ^14,16^. CLiX was run in default mode with ‘acronyms on’.

### MPSE

The Mendelian phenotype Search Engine (MPSE) employs Human Phenotype Ontology (HPO) based descriptions to prioritize patients, determining the likelihood that a Mendelian condition underlies a patient’s phenotype, based upon a training dataset. MPSE does not attempt to determine which Mendelian disease might underlie the patient’s phenotype, rather it seeks to categorize patients as positive or negative for Mendelian disease. MPSE employs a simple, well-established approach: Naïve Bayes ^17^. Briefy, MPSE uses the differences in HPO term frequencies between a collection of cases and controls to score each proband. The algorithm employs the BernoulliNB package from *scikit-learn*, a general-purpose machine learning library written in the Python programming language ^18^. We also discovered that the number of terms in a proband’s HPO description correlated modestly with age (r2 = 0.0725), accordingly, we used a linear regression to control for this effect. Although one can envision many algorithmic approaches to classification other than Naïve Bayes, e.g., Support Vector Machines or Neural nets, for this proof of principle study, we sought to demonstrate feasibility and provide baseline performance metrics. Future work will explore more sophisticated approaches to data modeling.

### Cross validation

We validated our results using leave-one-out cross validation, (i.e., k-fold cross validation, with k = 1) ^19^. This is the most stringent form of cross validation ^20^. More specifically, using the RCHSD data, we created 1049 different training datasets—each differing by a single proband—scoring each proband against a (different) version of MPSE, trained using a data subset that did not contain the proband being scored. All performance metrics were computed using this cross-validation scheme. We also carried out a second independent replication study using the clinical notes of 2965 University of Utah Level-III NICU admits. This dataset includes 35 WGS probands sequenced to date by the University of Utah NeoSeq program ^15^.

## Results & Discussion

Previous work ^10,12,21^, including our own ^14^, has demonstrated the utility of HPO-based, CNLP-derived phenotype descriptions for post sequencing diagnostic applications. Here we explore the feasibility of using CNLP phenotype descriptions, manufactured using the same NLP protocols, for triaging patients for WGS. To do so, we combined a Natural Language Processing (NLP) workflow based around the commercially available CLiX tool ^16^ with an ML-based prioritization tool we call MPSE, the Mendelian Phenotype Search Engine.

MPSE (see Methods) employs the Human Phenotype Ontology (HPO) ^11^ to prioritize patients. The *a priori* likelihood that a patient has a Mendelian condition is a computed probability based on the existence of HPO terms in the patient’s phenotype that are similar to those patients who previously had WGS. To investigate feasibility, we utilized curated RCHSD clinical data: 1049 Level IV NICU admissions and their clinical notes. Of these 1049 patients, 293 had rWGS and 85 received a molecular diagnosis. We validated the results presented below using leave-one-out cross validation; see methods for details. To examine the broader applicability of the RCHSD training data to other NICUs, we also carried out a second independent replication study using the clinical notes of 2965 patients from the University of Utah Level III NICU.

### Automated generation of HPO terms

We obtained HPO phenotype descriptions for all probands from clinical notes using Clinithink, a third party NLP tool ^16^. Automatically generating phenotypic descriptions via NLP is a major strength, as it enables the creation of large and dynamic pools of HPO-based phenotype descriptions for downstream prioritization activities.

Comparison of the CNLP descriptions to their corresponding manually compiled ones revealed notable differences with regards to HPO term numbers and contents. The CLiX generated descriptions for the RCHSD and NeoSeq cohorts had an average of 114.8 terms (min: 3, median: 91, max: 1000) and 64.5 terms (min: 1, median: 58, max: 300) respectively, whereas the corresponding manually created descriptions averaged 4.1 terms (min: 1, median: 3, max: 24) and 9.5 terms (min: 3, median: 9, max: 16) respectively.

### Prioritizing patients

We first sought to evaluate how effective our CNLP/MPSE pipeline was at prioritizing patients for WGS. In other words, did the children originally selected for WGS by physicians have higher MPSE scores than those who were not selected? **Figure 1** demonstrates that this is the case. As can be seen, the distributions of MPSE raw scores for the RCHSD and Utah WGS-selected children are well-separated from unsequenced ones. RCHSD sequenced cases had an average MPSE score of 26.6 while unsequenced controls had an average score of -31.7, statistically different by Student’s independent samples t-test (p<2e-16). The difference in mean MPSE score between Utah sequenced cases (17.3) and unsequenced controls (−33.7) was also statistically different (p=2e-12). The insert shows a Receiver Operator Characteristic (ROC) curve for the RCHSD data (AUC 0.86), indicating that MPSE can effectively prioritize probands for rWGS. The corresponding AUC for the Utah data was 0.85, essentially identical to the RCHSD result (ROC curve not shown).

**Figure 1.**
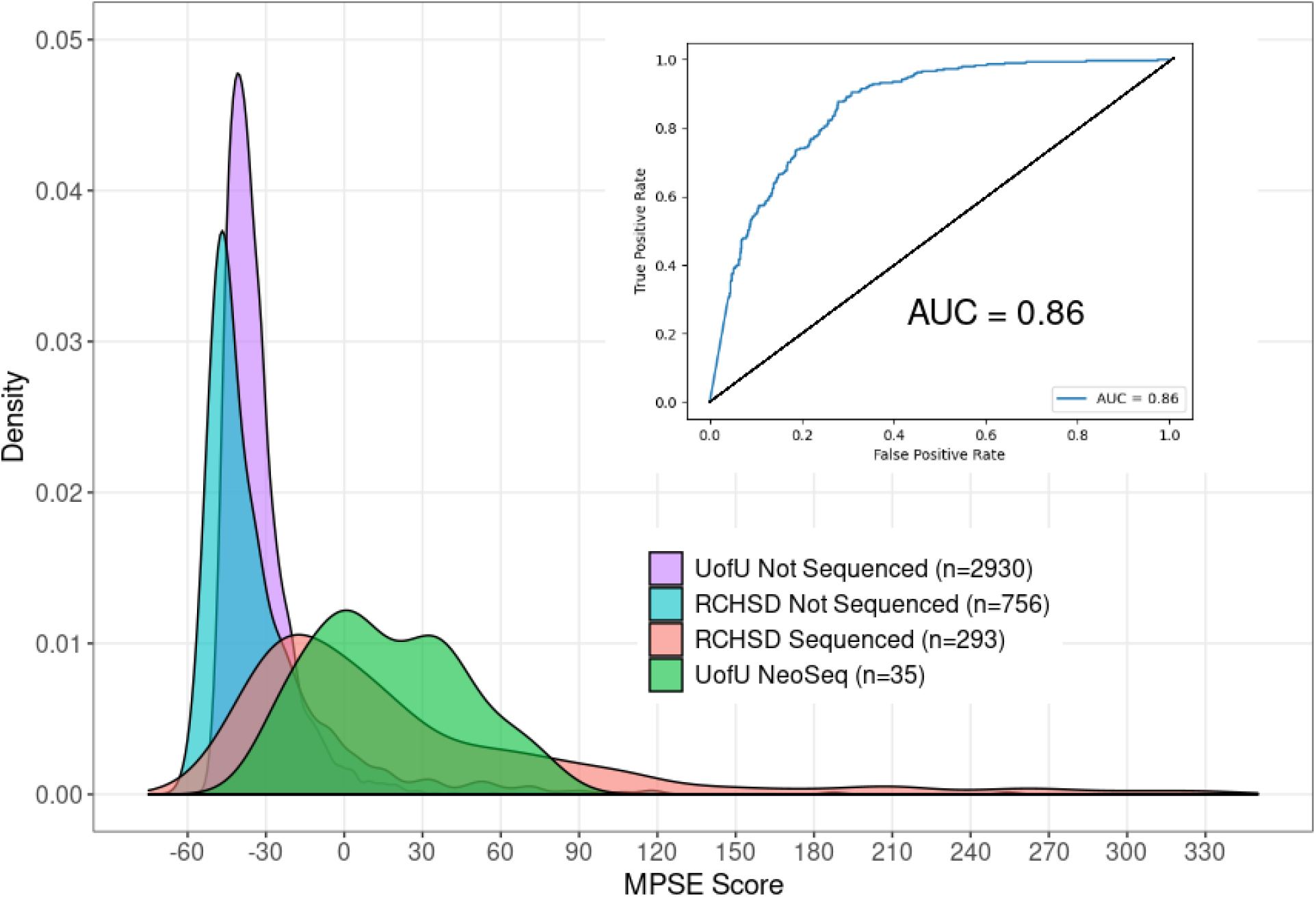
Automatically identifying probands with Mendelian phenotypes and prioritizing them for WGS using NLP-derived HPO phenotype descriptions. Panel A: distributions of MPSE raw scores for RCHSD sequenced (red), and RCHSD unsequenced (blue) probands. Score distributions for Utah NeoSeq (green) and Utah unsequenced probands (purple). Insert: Receiver Operator Characteristic (ROC) curve for RCHSD data. MPSE Scores are -log likelihood ratios.

### Cardinal phenotype terms

MPSE also provides means to identify, and highlight for expert review, those terms in a phenotype description that are most consistent with Mendelian disease. We refer to these terms as the proband’s cardinal phenotypes. **Figure 2** shows a CNLP phenotype description as a word cloud, wherein font sizes have been scaled by their individual contributions to the proband’s final MPSE score; those with the highest scores are shown in red, these are the proband’s MPSE cardinal phenotypes. These views of the patient’s phenotype description are designed to speed physician review and improve explainability.

**Figure 2.**
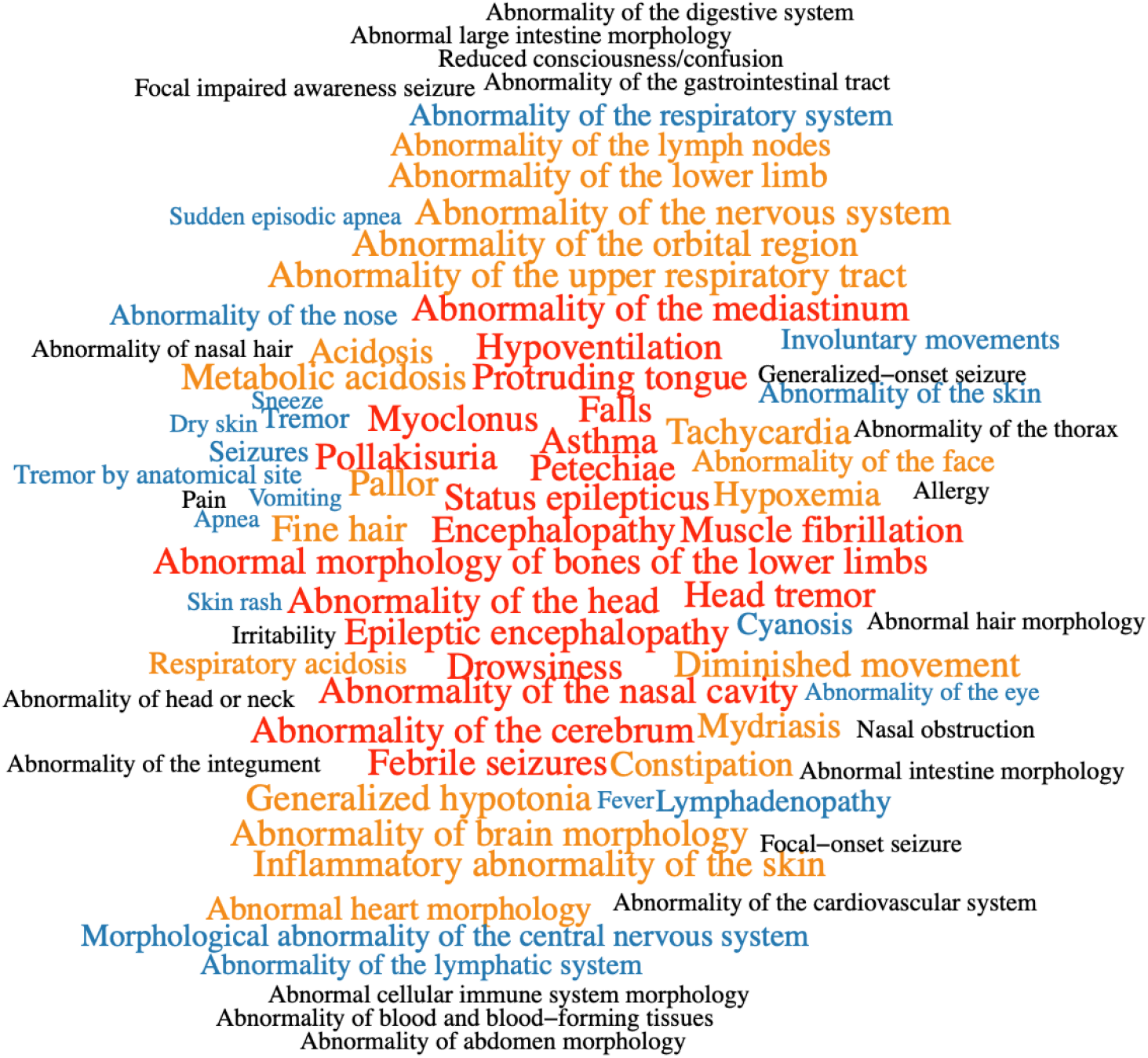
An automatically generated HPO-based phenotype description scored by MPSE. In this word-cloud, size and color are proportional to each HPO term’s contribution to the proband’s final MPSE prioritization score. Previously diagnosed by RCHSD using WGS, this child is heterozygous for a large deletion on the X chromosome which spans the PCDH19 gene, causative for female-restricted X-linked Epileptic Encephalopathy.

### MPSE diagnostic rates

To estimate MPSE-driven diagnostic rates, RCHSD and University of Utah sequenced probands were scored using leave-one-out cross validation, as described in Methods. The diagnostic fraction for these cohorts was 29% (85/293) and 43% (15/35), respectively. It should be borne in mind that this RCHSD diagnostic rate is for the specific dataset under analysis. It is not the RCHSD institutional WGS diagnostic rate. To facilitate comparison between these groups, we randomly re-sampled the larger RCHSD dataset so that it too had a 43% (85/198) diagnostic rate.

**Figure 3** shows projected diagnostic rates for these cohorts as a function of their MPSE scores. The negative slopes of the red, green and blue curves indicate that when using CNLP, higher MPSE scores are associated with diagnosed probands at both institutions. For instance, the top 25% of probands ranked on their MPSE scores from CNLP-generated phenotypes show very high diagnostic rates, approaching 100% for the highest MPSE scores. Moreover, for the CNLP datasets, diagnostic rates remain at or above the cohort diagnostic fraction of 43% at every MPSE score percentile. In contrast, the MPSE scores calculated from manually curated phenotypes (gray curve) are at best weakly associated with diagnostic status. This is not a result of inferiority of the physician-generated phenotypes, rather it is due to the fact that MPSE was trained using deep CNLP-derived phenotype data; recall that CNLP compared to manual review resulted in 64.5 vs 9.5 HPO terms/proband, respectively. Collectively, these results indicate that an MPSE-based prioritization pipeline could increase diagnostic rates above those obtained through expert manual case-review.

**Figure 3.**
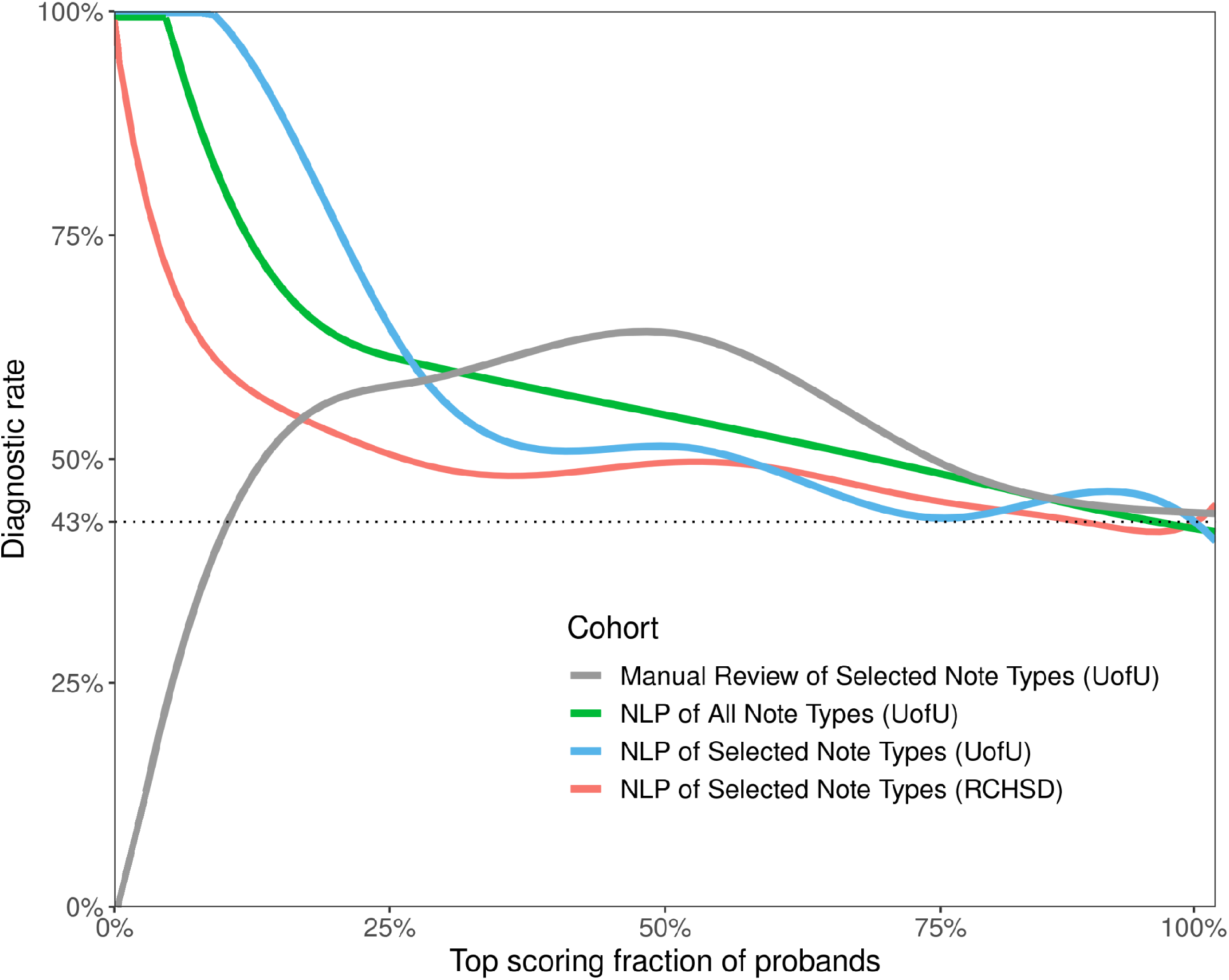
MPSE projected diagnostic rates. Higher MPSE scores correspond to increased probability of diagnosis, and projected diagnostic rates remain at or above the cohort diagnostic fraction of 43% at every MPSE score percentile.

### Impact of Note Types

Both RCHSD and the U of Utah limit manual review of clinical notes to a subset of note types deemed most informative by their institution’s expert reviewers. This is done to speed review by avoiding less informative and redundant note types. A potential advantage of CNLP is that volume is no longer an issue, and every note can be processed. We thus sought to evaluate the utility of processing all notes for every proband. The results of this experiment are also shown in **Figure 3**, where the blue and green curves summarize diagnostic enrichment as a function of MPSE score and note volumes. AUC for the top 50% of high scoring probands using all clinical notes vs. using only the selected note types is quite similar—62% and 65%, respectively. Thus, for the Utah dataset, using all available notes for every proband does not negatively impact diagnostic rates.

### Impact of Patient Populations

It is worth noting that underlying NICU populations differ between RCHSD and the University of Utah. Whereas RCHSD is a Level IV NICU, the University of Utah operates a Level III NICU, with the most severely ill patients transferred to Intermountain’s Primary Children’s neighboring Level IV facility. Thus, patients in the Utah dataset are likely to have fewer conditions requiring surgical interventions and a higher level of intensive care. Despite being trained using the RCHSD Level IV data, **Figure 1** makes it clear that the lesser acuity of Level III patients compared to Level IV patients did not interfere with MPSE’s ability to identify suitable candidates for sequencing. Nor did it negatively impact the correlation between MPSE score and Mendelian diagnostic rates (**Figure 3**). This finding suggests MPSE’s robustness to differences in NICU patient populations.

## Conclusions

We have demonstrated the feasibility of prioritizing individuals for WGS, using solely automated means, and that an automated process can meet or exceed diagnostic yields obtained through manual review of clinical notes. More sophisticated machine learning techniques might further improve performance. Neural and Bayesian networks, and Random Forest-based approaches generally outperform naïve Bayes. Likewise, addition of other metadata such as provider billing codes, medication histories, ancestry and socioeconomic indicators might still further improve performance. Nevertheless, even without such enhancements, our CNLP/MPSE workflow prioritized patients for rWGS with relatively high accuracy (AUC=0.86), with maximal projected diagnostic yields highly enriched for the top scoring quartile. These results bode well for future improved versions of the pipeline.

The ability of MPSE to accurately distinguish sequenced from unsequenced probands at both RCHSD and the University of Utah demonstrates the generalizability of the RCHSD training data, at least between two leading research institutions. The fact that MPSE was trained using RCHSD’s Level IV NICU patients, and replicated in Utah’s Level III NICU also provides some indication of MPSE’s robustness and applicability. Broader generalization, however, remains to be proven. Generalization is important because as WGS-based diagnosis becomes more widespread, and patients considered for testing become more diverse, clinical cultures and institutional differences in clinical note taking might render the parameters derived from the RCHSD training dataset less effective at some sites. In this regard, the ability of the pipeline to consume all notes for every proband is clearly an advantage, as it means adopters need not establish cross institutional equivalents in note types; instead, they can simply harvest every available clinical note for every proband.

More broadly, generalizability of training data must be distinguished from generalizability of the CNLP/MPSE workflow. The CNLP portion of the pipeline can be used to create a similar dataset for any institution engaged in WGS-based diagnosis, and, because it is a bayesian classifier, retraining MPSE using these data is straightforward. While we chose to use the CLiX CNLP tool, any NLP software able to produce high-fidelity HPO-based phenotype descriptions could be used upstream of MPSE. Going forward, we will explore the utility of retraining and combining models derived from multi-institutional datasets to further improve performance.

Recent work has also demonstrated the utility of WGS for Pediatric Intensive Care Unit (PICU) patients, where genome-based diagnoses have ended years-long diagnostic odysseys ^22^. The PICU generally has a more heterogeneous patient population than the NICU, because it includes patients from less than 12 months through 18 years of age, and a broader array of medical conditions such as cancer, organ transplant, trauma, etc. Thus, an automated tool such as MPSE that could help identify the relatively less common percentage of PICU patients with underlying Mendelian disorders could be especially useful for this population. These facts suggest that large medical systems may have other, non-pediatric patients who would also benefit from WGS—if they could be found. MPSE could in principle be used to search Electronic Medical Record databases for such patients. Outpatient pediatric specialty clinics might also benefit from using this type of automated tool.

Re-analysis of previously negative WGS cases is also increasingly an issue. The last decade has witnessed a huge increase in numbers of genes and variants associated with Mendelian conditions ^23,24^, with 250 newly described disorders annually, suggesting that many individuals previously undiagnosed by gene panels, WES, and WGS, could benefit from reanalysis in light of our ever-expanding knowledge of genetic disease. Recent work has validated this hypothesis ^25,26^. However, limited reimbursement and resources mean that, to be cost-effective, only those patients with the highest likelihood of diagnosis are currently reanalyzed using WGS technologies. Once again, automated approaches such as the one described here, might provide a means to locate and prioritize these patients for reanalysis. High MPSE scores might also be used to strengthen arguments for reimbursement. More generally, we foresee MPSE as an electronic decision support tool for facilitating the patient review process.

## Data Availability

All data produced in the present study are available upon reasonable request to the authors.

## Acknowledgments

We thank Aaron Quinlan and Gabor Marth for many helpful discussions in the early stages of the project.

## Funding Acknowledgment

The preparation of this manuscript was supported by a grant from the Conrad Prebys Foundation.

## Author Contributions

MY, BP, CH, and SK wrote the manuscript. BP, JH MY, CH, SK, SMJ, and LB designed the study and analysis strategy. MY, BP, and JH developed the MPSE algorithm. CH, SMB, and MY guided requirements. MM, BP, JH, SZ, and MY performed data analysis. EJ, ESK, and MNB compiled cases and clinical evidence. CH provided feedback on features and development. AO, SMJ, LB, and SK sponsored the project and provided helpful discussions and edits of the manuscript.

## Ethics approval and consent to participate

The need for Institutional Review Board Approval at Rady Children’s Hospital for the current study was waived as all data used from this project had previously been generated as part of IRB approved studies and none of the results reported in this manuscript can be used to identify individual patients. The studies from which cases were derived were previously approved by the Institutional Review Boards of Rady Children’s Hospital. The University of Utah Institutional Review Board approved the use of human subjects for this research, under a waiver for the requirement to obtain informed consent.

## References

1. Church G. Compelling Reasons for Repairing Human Germlines. N Engl J Med. 2017;377(20):1909–1911. doi:10.1056/NEJMp1710370

2. Farnaes L, Hildreth A, Sweeney NM, et al. Rapid whole-genome sequencing decreases infant morbidity and cost of hospitalization. NPJ Genomic Med. 2018;3:10. doi:10.1038/s41525-018-0049-4

3. Petrikin JE, Cakici JA, Clark MM, et al. The NSIGHT1-randomized controlled trial: rapid whole-genome sequencing for accelerated etiologic diagnosis in critically ill infants. NPJ Genomic Med. 2018;3:6. doi:10.1038/s41525-018-0045-8

4. French CE, Delon I, Dolling H, et al. Whole genome sequencing reveals that genetic conditions are frequent in intensively ill children. Intensive Care Med. 2019;45(5):627–636. doi:10.1007/s00134-019-05552-x

5. Kingsmore SF, Cakici JA, Clark MM, et al. A Randomized, Controlled Trial of the Analytic and Diagnostic Performance of Singleton and Trio, Rapid Genome and Exome Sequencing in Ill Infants. Am J Hum Genet. 2019;105(4):719–733. doi:10.1016/j.ajhg.2019.08.009

6. Dimmock D, Caylor S, Waldman B, et al. Project Baby Bear: Rapid precision care incorporating rWGS in 5 California children’s hospitals demonstrates improved clinical outcomes and reduced costs of care. Am J Hum Genet. 2021;108(7):1231–1238. doi:10.1016/j.ajhg.2021.05.008

7. Smedley D, Robinson PN. Phenotype-driven strategies for exome prioritization of human Mendelian disease genes. Genome Med. 2015;7(1):81. doi:10.1186/s13073-015-0199-2

8. Singleton MV, Guthery SL, Voelkerding KV, et al. Phevor combines multiple biomedical ontologies for accurate identification of disease-causing alleles in single individuals and small nuclear families. Am J Hum Genet. 2014;94(4):599–610. doi:10.1016/j.ajhg.2014.03.010

9. Cipriani V, Pontikos N, Arno G, et al. An Improved Phenotype-Driven Tool for Rare Mendelian Variant Prioritization: Benchmarking Exomiser on Real Patient Whole-Exome Data. Genes. 2020;11(4). doi:10.3390/genes11040460

10. Birgmeier J, Haeussler M, Deisseroth CA, et al. AMELIE speeds Mendelian diagnosis by matching patient phenotype and genotype to primary literature. Sci Transl Med. 2020;12(544):eaau9113. doi:10.1126/scitranslmed.aau9113

11. Groza T, Köhler S, Moldenhauer D, et al. The Human Phenotype Ontology: Semantic Unification of Common and Rare Disease. Am J Hum Genet. 2015;97(1):111–124. doi:10.1016/j.ajhg.2015.05.020

12. Clark MM, Hildreth A, Batalov S, et al. Diagnosis of genetic diseases in seriously ill children by rapid whole-genome sequencing and automated phenotyping and interpretation. Sci Transl Med. 2019;11(489):eaat6177. doi:10.1126/scitranslmed.aat6177

13. James KN, Clark MM, Camp B, et al. Partially automated whole-genome sequencing reanalysis of previously undiagnosed pediatric patients can efficiently yield new diagnoses. Npj Genomic Med. 2020;5(1):1–8. doi:10.1038/s41525-020-00140-1

14. De La Vega FM, Chowdhury S, Moore B, et al. Artificial intelligence enables comprehensive genome interpretation and nomination of candidate diagnoses for rare genetic diseases. Genome Med. 2021;13(1):153. doi:10.1186/s13073-021-00965-0

15. Nicholas TJ, Al-Sweel N, Farrell A, et al. Comprehensive variant calling from whole-genome sequencing identifies a complex inversion that disrupts ZFPM2 in familial congenital diaphragmatic hernia. Mol Genet Genomic Med. 2022;10(4):e1888. doi:10.1002/mgg3.1888

16. Clinithink. Clinithink: AI Solutions Company, Clinical Data Solutions for Life Science & Healthcare. Accessed March 5, 2021. https://www.clinithink.com

17. Ng AY, Jordan MI. On Discriminative vs. Generative Classifiers: A comparison of logistic regression and naive Bayes. :8.

18. Pedregosa F, Varoquaux G, Gramfort A, et al. Scikit-learn: Machine Learning in Python. J Mach Learn Res. 2011;12(85):2825–2830.

19. Hastie T, Friedman J, Tibshirani R. The Elements of Statistical Learning. 1st ed. Springer New York, NY; 2001. Accessed April 20, 2022. https://link.springer.com/book/10.1007/978-0-387-21606-5

20. Molinaro AM, Simon R, Pfeiffer RM. Prediction error estimation: a comparison of resampling methods. Bioinforma Oxf Engl. 2005;21(15):3301–3307. doi:10.1093/bioinformatics/bti499

21. Deisseroth CA, Birgmeier J, Bodle EE, et al. ClinPhen extracts and prioritizes patient phenotypes directly from medical records to expedite genetic disease diagnosis. Genet Med. 2019;21(7):1585–1593. doi:10.1038/s41436-018-0381-1

22. Sanford EF, Clark MM, Farnaes L, et al. Rapid Whole Genome Sequencing Has Clinical Utility in Children in the PICU. Pediatr Crit Care Med J Soc Crit Care Med World Fed Pediatr Intensive Crit Care Soc. 2019;20(11):1007–1020. doi:10.1097/PCC.0000000000002056

23. Bamshad MJ, Nickerson DA, Chong JX. Mendelian Gene Discovery: Fast and Furious with No End in Sight. Am J Hum Genet. 2019;105(3):448–455. doi:10.1016/j.ajhg.2019.07.011

24. Amberger JS, Bocchini CA, Schiettecatte F, Scott AF, Hamosh A. OMIM.org: Online Mendelian Inheritance in Man (OMIM®), an online catalog of human genes and genetic disorders. Nucleic Acids Res. 2015;43(Database issue):D789–798. doi:10.1093/nar/gku1205

25. Liu P, Meng L, Normand EA, et al. Reanalysis of Clinical Exome Sequencing Data. N Engl J Med. 2019;380(25):2478–2480. doi:10.1056/NEJMc1812033

26. Wenger AM, Guturu H, Bernstein JA, Bejerano G. Systematic reanalysis of clinical exome data yields additional diagnoses: implications for providers. Genet Med Off J Am Coll Med Genet. 2017;19(2):209–214. doi:10.1038/gim.2016.88

